# Multidimensional Bayesian Estimation for Deep Brain Stimulation Using the SafeOpt Algorithm

**DOI:** 10.1101/2022.01.30.22270042

**Authors:** Scott E. Cooper, Théoden I. Netoff

## Abstract

Some symptoms treated with Deep Brain Stimulation (DBS) such as gait in Parkinson’s disease (PD), are often poorly responsive to DBS. This may be because DBS settings are usually optimized to other symptoms. To test this, we require an efficient, safe optimization algorithm. To develop such a tool, we extend the BayesOpt algorithm whose successful application to DBS settings we previously published [Louie et al 2021 *J Neuroeng Rehabil*], using, as a test bed, a simulated cost function constructed for biological plausibility, with measurement noise based on experimental data.

We found that the SafeOpt algorithm [Sui et al 2015 *Proc Machine Learning Res*] converged to the optimum as well, and as fast as the BayesOpt algorithm, while avoiding high-cost points much more effectively. In three dimensions, SafeOpt converged in about 30 iterations, which is a feasible number for physical experiments in real patients. Convergence was slower when measurement nose was greater, but this could be overcome by running it for more iterations. The algorithm was relatively robust to misspecification of hyperparameters, and considerably more robust when hyperparameter fitting was incorporated into the algorithm. The algorithm did not perform as well when the quantization of stimulation settings was coarser, suggesting that it will work better with neurostimulators capable of independent current control. Finally, the algorithm was able to cope with a cost function having multiple local minima.

## 1 Introduction

### 1.1 Challanges in clinical DBS optimization

Deep brain stimulation is a common and effective treatment for Parkinson’s disease^*1–4*^ and other neurological disorders^*5, 6*^. Once the lead is surgically implanted in the brain, the electrical stimulation parameters must be adjusted to suit the individual patient, in order to obtain satisfactory clinical benefit. The current state of the art is for a skilled expert clinician to set the stimulation parameters, stimulate, assess symptoms, change the parameters, assess again, and so on, until satisfactory results are achieved^*7–9*^.

This has obvious drawbacks. First, it depends on skilled experts who may be in short supply, particularly in rural areas and medically underserved populations. Second, human clinicians may exhibit bias and variability. Third, there is wide variation among clinicians in their approach to stimulation adjustments; this thwarts standardization in clinical trials, and hampers health-system-wide efforts at outcomes assessment and quality improvement.

Another drawback stems from the multidimensional nature of the adjustment process. This is increased with newer DBS technology incorporating directional leads and independent current control^*10*^, thereby increasing the dimensionality of the parameter space which the clinician must search, in order to find optimal, or even satisfactory results. Multidimensional optimization is challanging, for human clinicians, presenting an obvious opportunity for computer algorithms. In practice, clinicians typically rely on dimensionality-reducing heuristics, e.g. varying one parameter at a time^*7–9*^. If changes in parameters are small, and result in small changes in the outcome, then this works as gradient descent. However gradient descent can be inefficient when clinical time is of the essence for both patient and physician, and may only find only local minima without converging on a global minimum.

### 1.2 Bayesian estimation of a gausian process

A data-efficient alternative to gradient descent is based on estimating the cost function, rather than the optimum per se. This is the approach of Gaussian process estimation (GPE). GPE incoprates an explicit, quantitatively smoothness assumption via a kernel function; this allows points distant from the optimum to contribute information about the optimum, reducing noise. GPE also allows for quantification of uncertainty. GPE adapts easily to the clinical assess-adjust-assess cycle: at every i_th_ iteration the algorithm can choose the most informative i+1_th_ test point using the estimate of the cost function from the i points tested so far. This is Bayesian estimation, and it allows for a precise definition of “most informative” test point using criteria such as uncertainty-reduction, or likelihood of finding a better optimum.

We have previously published a proof of concept study in which we used the BayesOpt Bayesian estimation algorithm successfully for one-dimensional optimization of DBS treatment of rigidity in people with Parkinson’s disease^*11*^. This proof of concept can be extended to more dimensions.

### 1.3 Safety in optimization

BayesOpt learns the optimal setting efficiently over a set of bounded conditions. The bounds may include a priori safety limits, but the algorithm cannot learn safety limits from data. When the algorithm chooses the next set of stimulation parameters to be tested, there is always a risk that these parameters will be “bad,” not merely in the sense of failing to improve symptoms, but even worsening symptoms, or causing actual discomfort, or other side effects. Bayesian estimation needs to explore previously unexplored regions of parameter space, where minima (optima) may be “hiding,” but such regions can also have high risk of bad settings. In our Parkinson’s rigidity study, if a particular settng caused discomfort to the patient, then a human clinician changed the parameter bounds to prevent further testing of settings that were likely to be uncomfortable. This was feasible, in the one-dimensional case, with a parameter where we could assume a monotonic relation with side effects. For mor general applications, it is obviously desirable to build safety into the algorithm, rather than rely solely on a human clinician.

### 1.4 SafeOpt

The “SafeOpt” algorithm^*12, 13*^ is a Bayesian estimation method which starts with a single known-safe test point, and from there expands the safe region, adding each new point where it gives the most information (high uncertainty), subject to the requirement that probability of a very bad result must be low (upper bound of the confidence-inerval of the estimated cost function must be less than a safety threshold). As a result, it tends to explore outward from a safe region, towards a minimum, expanding the explored region around that minimum. The result is a cost function estimate that is accurate in the vicinity of the minimum, giving an accurate estimate of the true minimum.

### 1.5 Aim of this paper

We developed a SafeOpt implementation with the intention of testing it on DBS for gait in people with Parkinson’s disease. In particular, we needed to estimate how many iterations are required to find an optimum, and how robust this peformance is, in order to assess feasibility of experiments in humans. Simulations can give accurate answers to those questions only if the model cost function is biologically realistic, and on whether a realistic amount of measurement noise is incorporated in the model. Therefore, we devote careful attention to these issues below. Our goal was to learn as much as posible from the simulation in preparation for human experiments.

## 2 Methods

### 2.1 Software platform

Simulations were performed in the Julia programming language^*14*^ because example code for SafeOpt, was available in Julia^*13*^. Julia is a high-level language, optimized for matrix and array manipulation, similar to Matlab and Python/Numpy.

### 2.2 Model cost function

#### 2.2.1 DBS settings

In this paper, we adapt the SafeOpt algorithm to the practical constraints of an industry-standard “1331” (“directional”) DBS implantable lead and independent current control (“ICC”). We then tested it against a noisy cost function simulating the clinical response to varying parameters with such an electrode. We then used this model to assess accuracy of the estimated optimum, and the rate of convergance, and compared these to the BayesOpt algorithm, with particular attention to “safety,” i.e. did the algorithm choose very bad (high cost) test points. Finally, we assessed the robustness of SafeOpt accuracy and convergence to variation in measurement noise, granularity of the simulation settings, hyperparameter misspecification, and the characteristics of the cost function.

In this paper, we focus on the three-dimensional case, optimizing over 3, out of a possible 5 parameters. These are the three parameters which receive the most attention in clinical stimulator adjustment, because they determine where in the region brain surrounding the implanted lead stimulation has its effect. This is believed to be a critical determinant of clinical effect^*15*^. The two parameters not considered here are pulse width, and frequency; in clinical adjustment, these are often set heuristically to constant values, and not tuned unless tuning the other three fails to produce a satisfactory effect.

The three parameters considered here are: amplitude, level, and direction. Amplitude is simply the total electrical current delivered, and determines how far DBS effect spreads radially away from the electrode. Level is the position along the long axis of the DBS lead where stimulation is delivered. This is adjustable because DBS leads have multiple contacts positioned along their length. Current can be delivered to two adjacent contacts simultaneously, giving a level effectively intermediate between the two. With independent current control (ICC)^*16*^, the proportion of current delivered to each contact is controlled, allowing the intermediate level to vary continuously from 100% one contact to 100% the other. Direction is adjustable because contacts at a given level are split into three segments 120 degrees apart. Here, too, intermediate stimulation is possible by stimulating two segments, and ICC allows this to vary continuously. The three parameters can be thought of as coordinates in a cylindrical coordinate system: rho = amplitude, theta = direction, z = level. Existing neurostimulators discretize these “continuous” quantities The SafeOpt algorithm is a natural fit for discretized parameters, because, by design it works with a finite feasible set of test points, rather than continuous quantities.

#### 2.2.2 Level and Direction parameters

For level, we employ a nomenclature based on Boston Scientific devices. Level is indicated as L1.0 … L4.0 numbering from the contact at he distal tip of the DBS lead. Intermediate levels are indicated as L1.1, L1.2, etc. L1.2, for example means 80% of the current goes to L1, and 20% to L2 resulting in a nominal level at 20% of the distance from L1 to L2. Direction is indicated in degrees, assigning 0 degrees to the anterior-facing segment (this is the segment aligned with the radiographic fiducial), and the others at 120 and 240 degrees.

Among DBS leads available for use in humans, all DBS leads with directional capability have 4 levels. All human DBS leads with directional capability have segmented leads only at levels 2 & 3. (Older devices from Medtronic and Boston Scientific lack directional capability.) This means that the direction parameter is undefined at L1.0 and L4.0, and we therefore omit them from the feasible set, although L1.1 and L3.9 are included.

#### 2.2.3 Scaling of DBS parameters

The three parameters are measured in different units. We could account for these differences by complicating the kernel of the Gaussian process, but we have taken a simpler, more pragmatic approach: we use an isotropic kernel, and scale all 3 parameters to a common zero to 1.0 range. This is possible because two parameters (level and direction,) have natural upper and lower limits, while the amplitude has a natural lower limit (zero), and a practical upper limit (amplitudes above 6 mA are rare, due to high battery consumption and, often, side effects, as well as concerns about tissue damage at high current density^*17*^).

#### 2.2.4 Cost function

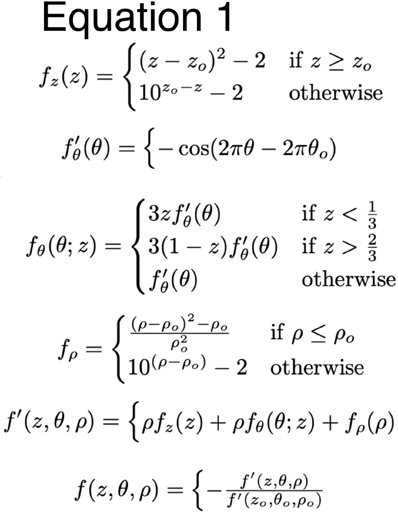

We tested the algorithm against a model cost function designed for biological plausibility. This allowed us to validate the algorithm against ground truth, and to manipulate the cost function in ways which stress-tested the algorithm. The cost function Equation 1 was constructed from three costs, reflecting, respectively, the influence of amplitude, level, and direction. The amplitude cost was zero at an amplitude of zero (i.e. no DBS), declined polynomially to a minimum of -1.0, then increased exponentially above that. The level cost had the same form as the amplitude cost, that is: polynomially decreasing from highest level down to a minimum of -1.0, and increasing exponentially at levels below that. The direction cost was a simple cosine, with one parameter specifying the location of the minimum. The cosine was “flattened” progressively as a function of level, such that direction had the most effect when level was between L2.0 and L3.0, which are segmented contacts, and no effect when level approached L1.0 and L4.0, since, as noted above, in this case, these contacts are unsegmented, and provide no directional current “steering.” The level and direction costs were multiplied by the amplitude to make them zero when amplitude was zero; this is to achieve the biologically obvious behavior that it doesn’t matter what the level or direction is, if the electric current amplitude is zero. The three terms were then summed and scaled so that the range of the cost function was [-1,+1] with a value of zero corresponding to no stimulation (zero amplitude). This modeled the (clinically common) case where DBS might worsen the target symptom; severe side effects would be represented with values in excess of +1. Measurement noise was added as Gaussian, with SD of 0.5 (experimentally determined, section 2.3), thus the optimum was 2 SD below the no-stimulation condition. In Figure 1 lower right, the cost function is shown as a colormap (blue: negative; yellow: positive). The *marginal* function values are shown, i.e. in each 2D plot, the value shown is averaged over all values of the omitted dimension.

**Figure 1.**
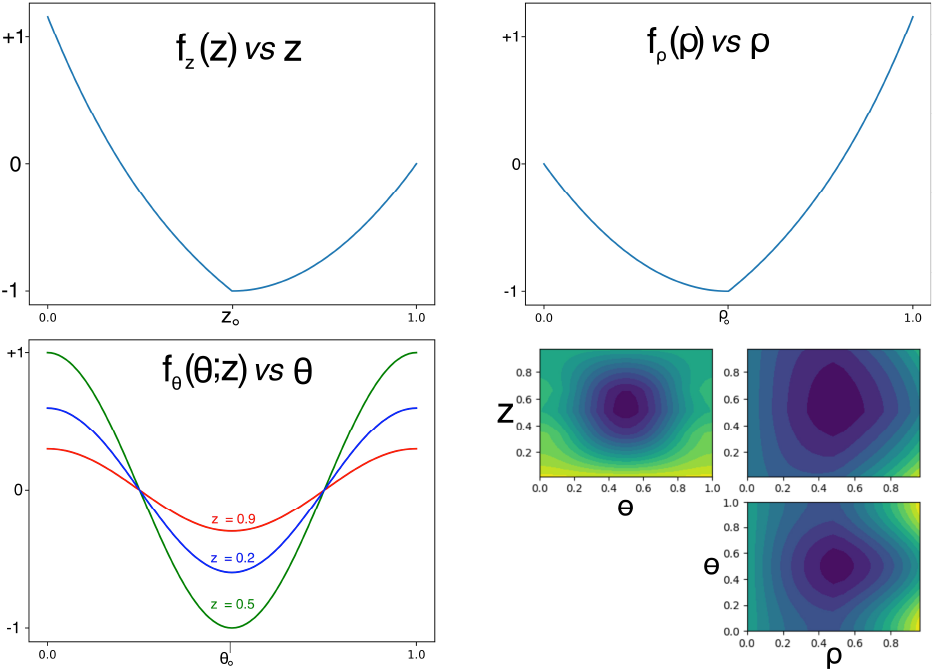

### 2.3 Experimental determination of measurement noise

To simulate measurement noise in a physical experiment Gaussian noise was added to the cost function. In our experimental work we focus on a particular disease symptom, namely gait impairment in Parkinson’s disease. Therefore, we obtained an estimate of measurement noise, relative to DBS effect size, from experimental gait data, based on step and stride length. First, we used data from a published experiment^*18*^, in which we measured step length in 27 Parkinson’s patients, during one minute of walking, repeated the measurement 5 times, and, from those 5 trials computed the standard deviation, dividing by the average to give the standard deviation represented as a percent change, for each subject. The median over all 27 patients was 1.8%,1.6% for the left and right foot, respectively. Next, from our previously published study of DBS effects^*19*^ we obtained an effect size for the therapeutic effect of DBS on stride length, using a similar one-minute walking protocol. As Figure 1E in that paper shows, the effect size was about 5%, i.e. the therapeutic effect of DBS was a 5% increase in stride length. Note that stride length is the sum of left plus right step length, so we take 5% as the therapeutic effect of DBS on step length. This is a conservative estimate of DBS gait effect size, because measurements were made immediately after adjusting DBS settings, without waiting for full wash-in. We thus can estimate that the measurement noise SD, relative to effect size is about 1.7/5 or 0.34. The effect size in our cost function is the difference between optimal stimulation (−1.0) and no stimulation (0.0), so we could set noise SD at 0.34, however, we chose a more conservative value of 0.5.

### 2.4 SafeOpt implementation

#### 2.4.1 Gaussian process

A Gaussian process generate a Gaussian probability distribution, characerized by mean Ŷ_i_ and variance *ν*_i_ for the value of the unknown function at a test input **x**_**i**_ given a mean function m(**x**) and a covariance, or kernel function **k(x**,**x’)** and a set of previously tested inputs **X** with associated function outputs **y**.

#### 2.4.2 Kernel function, mean function

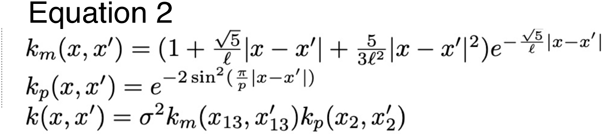

A kernel measures the distance, or dissimilarity between two points **x**,**x’** We used a kernel which was periodic on the *direction* axis and Matern 5/3 on the *level* and *amplitude* axes. The kernel **k(x**,**x’)** is the product of a Matern kernel **k**_**m**_**(x**_**13**_,**x’**_**13**_) and a periodic kernel **k**_**p**_**(x**_**2**_,**x’**_**2**_), where **x**=[*x*_*1*_,*x*_*2*_,*x*_*3*_], **x**_**13**_=[*x*_*1*_,*x*_*3*_], and **x**_**2**_=[*x*_*2*_]. In Equation 2 |.| denotes the norm (length) of a vector. Sigma is the signal standard deviation, l is the length constant, p is the period (1.0 in our case).

#### 2.4.3 Acquisition function

SafeOpt (like BayesOpt) incorporates function evaluations (measurements) at previous test points **X**, into a Gaussian process (GP), and uses the predictions to choose the best next test point **x**_i_ at which to make a measurement. Given a possible test point **x**, GP generates Ŷ and ν for that **x**, and the best next test point is the one which maximizes some acquisition function of Ŷ and *ν*. SafeOpt uses an “uncertainty” acquisition function, that is, it maximizes *ν*. Most of the power of the algorithm comes not from the acquisition function, but from the way SafeOpt defines, and refines the feasible set.

#### 2.4.4 Defining and refining the feasible set

SafeOpt starts with a finite set of potential test points. At each iteration, the GP is updated, and potential test points are classified into subsets based on the bounds of the confidence interval (CI). The CI is determined from Ŷ and *ν* as given by the GP. We use 98% CIs. ***S*** is the subset of *safe* points, defined as those for which the upper bound of the CI is less than a safety threshold Ymax (see section 4.2.5 below). ***M*** is the subset of ***S*** which are *potential minimizers* defined as points for which the lower bound of the CI is less than the Ŷ for any point tested so far. ***E*** is the subset of *expanders*, defined as points which, if tested, would increase the number of points in ***S***, if testing yielded a value at the lower bound of the CI. The intersection of ***M*** with ***E*** is then searched for the point which maximizes the acquisition function.

#### 2.4.5 Ymax

The threshold for defining safe points was set at +1.0, which is the upper limit where DBS worsens the target symptom, but without severe side effects. We found that, at the start of iteration, when few measurements had been made, and confidence intervals were, therefore, very wide, the set of safe points was sometimes empty. Therefore, we modified the algorithm so that, if the safe set was empty, Ymax was temporarily increased by 4/3, and if that failed, by 2. In practice, this rarely occured, and, when it did, Ymax only ever needed to be increased on the first or second iteration, and not thereafter.

#### 2.4.6 Initialization

Initializing the SafeOpt algorithm requires a single known-safe test point. In a practical application, this could come in either of two ways. In the first scenario, it could be the outcome of conventional optimization by a human clinician. In this case, the goal of the optimization is to improve the clinician’s result, or, if it is already optimal, increase confidence in its optimality. In the second scenario, there is no prior optimization. In this case, it makes sense to choose an initial point which may provide little therapeutic benefit (cost function *not very low*), but is probably “safe,” (cost function *not very high*). This corresponds to parameters where we know, a priori, that there will be no serious side effects, and no worsening of symptoms. This occurs when the amplitude is near zero. Also, for neuroanatomical reasons (distance from internal capsule ^*20, 21*^ and somatosensory thalamus^*22*^) there is usually an expected safe level furthest from the distal tip of the DBS lead (highest level), where side effects are least likely to ocurr. Therefore, the algorithm was initialized by evaluating the cost function for a single “known-safe” point with minimum amplitude, maximum level, and random direction.

#### 2.4.7 Hyperparameters

We used a *constant* mean function so the GP had four hyperparameters: the constant value returned by the mean function, the length scale, the signal variance, and the measurement noise variance. The function value from the first, “known-safe” point was used for the constant value of the mean function. Values for the three other hyperparameters were: length scale of 1.5, Noise SD 0.5 (i.e. the measurement noise) signal sd 3.0, and length scale 1.5.

The maximum possible distance between two points on each axis is (1, 0.5, 1) (because 0.5 is the furthest apart two points can be, around the circumference of a circle of diameter 1.0) therefore, the maximum possible distance between two points is sqrt(1^2^ + 0.5^2^ + 1^2^)=1.5 (Figure 2), making this a natural choice for the length constant.

**Figure 2.**
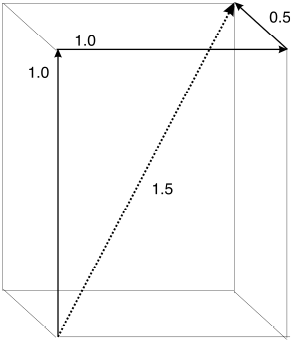

## 3 Results

### 3.1 SafeOpt behavior

Figure 3 shows the behavior of the SafeOpt algorithm optimizing over a three-dimensional cost function, simulating DBS effects on gait. <u>*Caption:*</u> The three-dimensional stim parameter space is shown as a set of three two-dimensional spaces. The true cost function is shown as a colormap (blue: negative; yellow: positive), while the Gaussian process estimate of the cost function is shown as a contour plot (dashed: negative; solid: positive). In both cases, the *marginal* function values are shown, i.e. in each 2D plot, the value shown is averaged over all values of the omitted dimension. The optimum of the GP estimate is shown as a red +. Test points are shown as white o. The initial “known safe” point is shown as white x.

**Figure 3.**
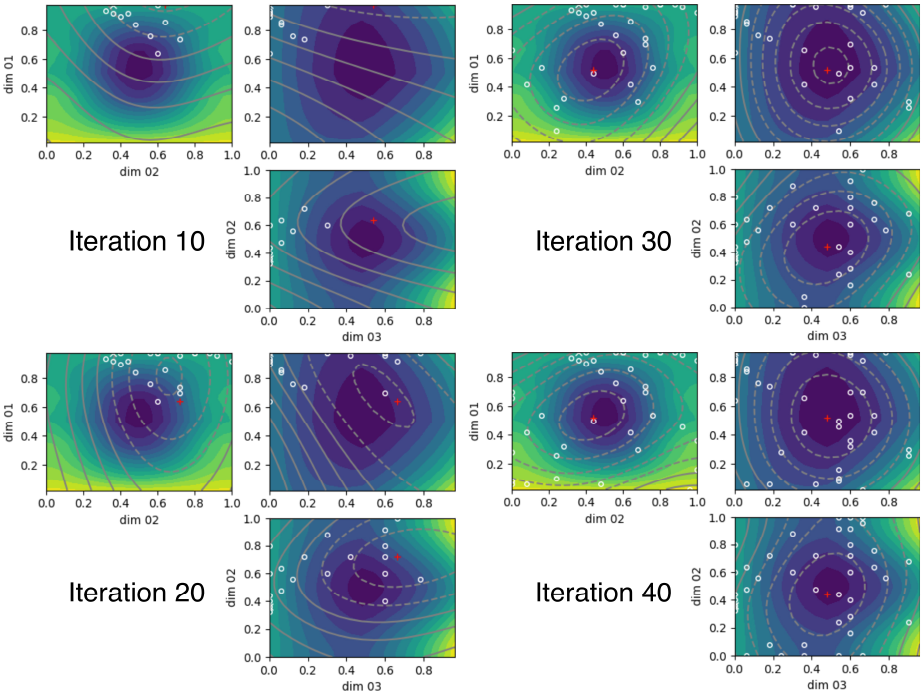

Figure 4 shows the SafeOpt algorithm converging within 30 iterations <u>*Caption:*</u> 16 runs of the SafeOpt algorithm. *Top left* shows the true cost of the estimated optimum converging towards -0.99 (the best possible value) with iteration number. *Bottom left* shows the distance from estimated to true optimum, converging towards zero with iteration number. In both cases, convergence is rapid, usually within 30 iterations On the right, the true cost of the estimated optimm for 64 runs is shown after 30 (*top right*) and 60 (*bottom right*) iterations.

**Figure 4.**
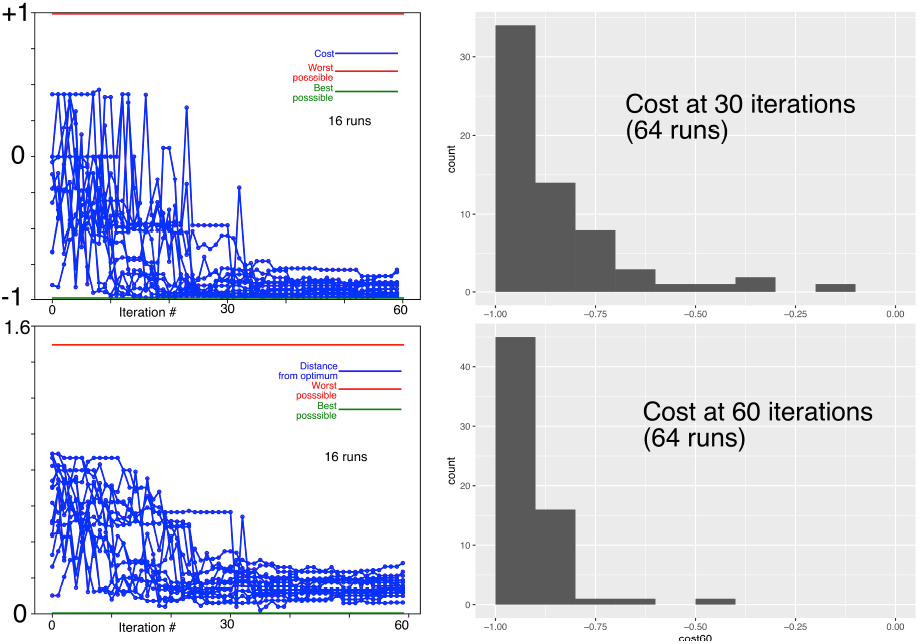

### 3.2 Comparison vs. BayesOpt

Figure 5 shows that SafeOpt and BayesOpt converge at similar rates, but BayesOpt explores bad (high cost) regions of stimulator parameter space very early (before convergence), whereas SafeOpt does not explore bad regions until late (after convergence). <u>*Caption:*</u> Comparison of SafeOpt with BayesOpt. *Top row* shows the true cost of the estimated optimum converging with iteration number. *Bottom row* shows that Bayes opt tests bad (high cost) test points well before convergance, while SafeOpt does not. The worst possible value of the cost function is +1.0, but due to the granularity of the feasible set, the highest cost in the feasible set is +0.96.

**Figure 5.**
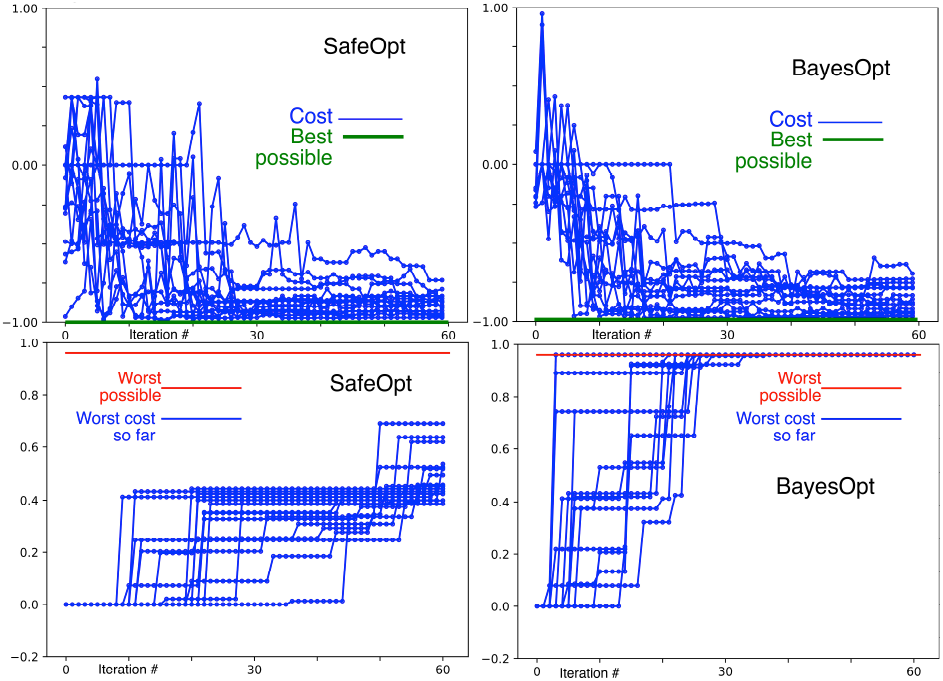

### 3.3 Sensitivity analysis

#### 3.3.1 Measurement noise

As noted above, we based simulated measurement noise on experimental gait data. To assess robustness of our results in the pessimistic case where measurements are noisier than expected, we doubled the measurement noise to 1.0. Figure 6 shows that convergance was slower, but still complete within 90-120 iterations. <u>*Caption:*</u> The true cost of the estimated optimum, at 30, 60, 90, and 120 iterations for 64 runs of the SafeOpt algorithm with measurement noise doubled. Convergance was slower, but still complete within 90-120 iterations.

**Figure 6.**
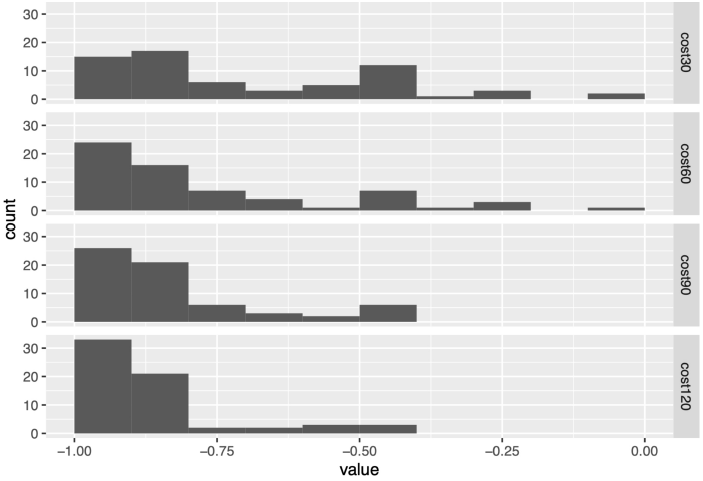

#### 3.3.2 Hyperparameters

We also explored the effect of a mismatch between the kernel hyperparameters of the GP, and and their best values. We simulated “grossly misspecified” values by perturbing each hyperparameter by a factor of two (one-half base value, and twice base value). This gave three values for each hyperparameter. We ran the algorithm for 60 iterations, 16 times for each of the 27 different combinations of values of hyperparameters. The algorithm failed to converge in 7/27=26% cases. When zero out of 3, or 1 out of 3 hyperparameters were misspecified, it *always* converged. When 2 out of 3 were misspecified, it failed in 4/12 cases (33%). When all 3 were misspecified, it failed in 3/8 cases (38%). In summary, the algorithm succeeds in a large majority of cases, even when 2, or even all 3 hyperparameters are misspecified, but it may fail if multiple hyperparameters are grossly (factor of 2) misspecified.

#### 3.3.3 Fitting hyperparameters

Hyperparameter misspecification can be improved upon. When there is uncertainty about the best value of a GP hyperparameter, a standard approach is to let the hyperparameters vary, by fitting them to the data, using maximum likelihood, or cross-validation. We implemented maximum likelihood fitting of the length constant *l*, signal standard deviation, and noise hyperparameters on every 5th iteration. We then applied this modification, with deliberately misspecified starting values of those hyperparameters, using values which reliably caused the unmodified algorithm to fail.

With this approach, convergence was much better (Figure 7). We have not pursued this approach further, in this paper, due to limitations of the Gaussian process software library we are using, but it appears promising for future implementation with different software. <u>*Caption:*</u> True cost of estimated optimum, versus iteration number, for grossly (factor of two) misspecified hyperparameters (*length*: 3.0, *signal SD* 1.5, *noise* 0.5), 16 runs, showing nonconvergence of the unmodified algorithm, and convergence with hyperparameters refit to the data at every 5th iteration.

**Figure 7.**
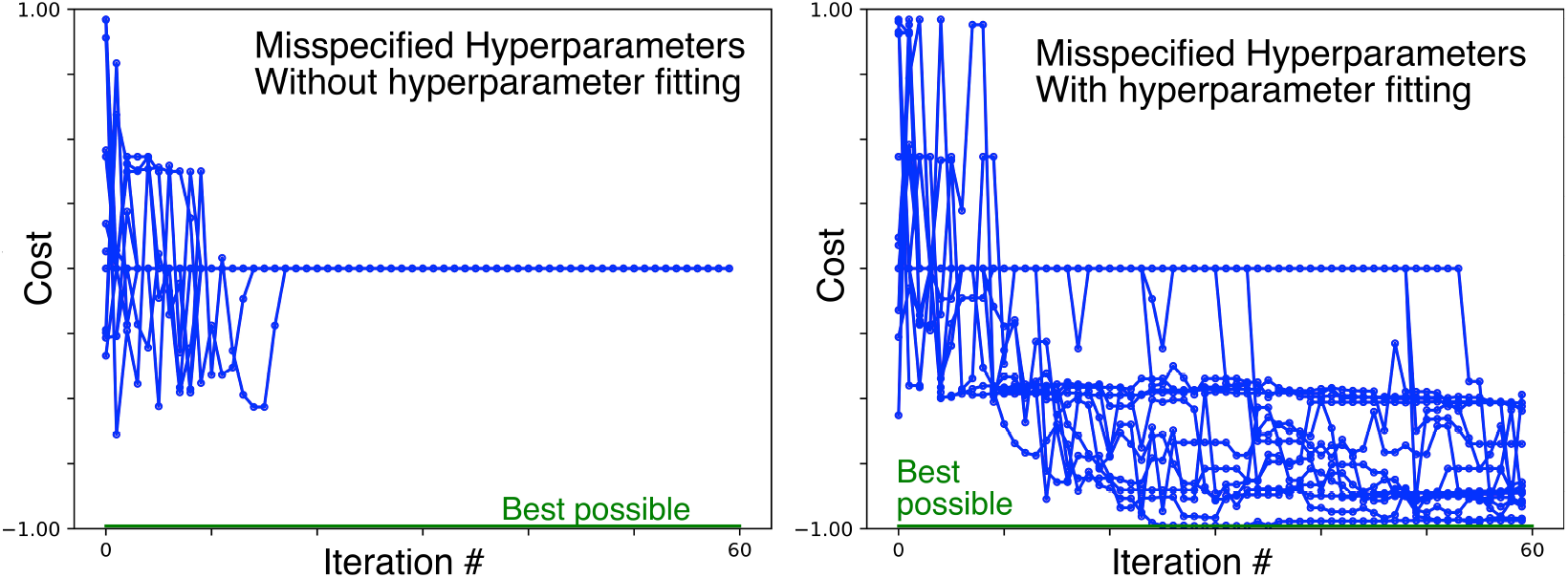

#### 3.3.4 Sensitivity to coarseness of feasible set

While in principle DBS stim settings are continuous quantities, in practice, implantable neurostimulators are digital devices which impose some granularity (smallest increment) on stim settings. As noted above SafeOpt handles this in a natural way. Granularity differs between different manufacturers’ devices. All manufacturers allow amplitude to be tuned in increments (0.1 mA or less) that are very small compared to minimum detectable clinical effect. For our normalized amplitude parameter, with range [0-1] we used an increment of 0.06 which, for an amplitude range of [0-6 mA] corresponds to 0.36 mA. However, granularity of the level and direction parameters differ significantly between manufacturers. Devices with independent current control allow these to be tuned in very small increments e.g. Boston Scientific allows increments of 10% of the distance between adjacent levels. In contrast the Abbott device lacks ICC and can approximate increments of 50% of the difference between adjacent levels (by applying the same voltage to adjacent contacts, although the proportion of current through each contact will depend on relative contact impedances).

Therefore, we examined the effect of coarsening the level and direction parameters; we increased the minimum increments from 0.02 (level) and 0.04 (for direction) to 0.15 (level) and 0.15 (direction). Mapping from our normalized stimulation parameters [0,1] to the physical DBS electrode (level [1-4], direction [0-2*π*] this corresponds approximately to the Abbott case. Results are shown in Figure 8. With this granularity, we found performance somewhat degraded, sometimes achieving only about 50% of the true optimum. This suggests that methods like SafeOpt will work less well for neurostimulators without ICC than for those with ICC. <u>*Caption:*</u> True cost of estimated optimum, versus iteration number, for the standard case, and for coarse granularity of direction and level settings, simulating a neurostimulator without independent current control (see text, section 3.3.4).

**Figure 8.**
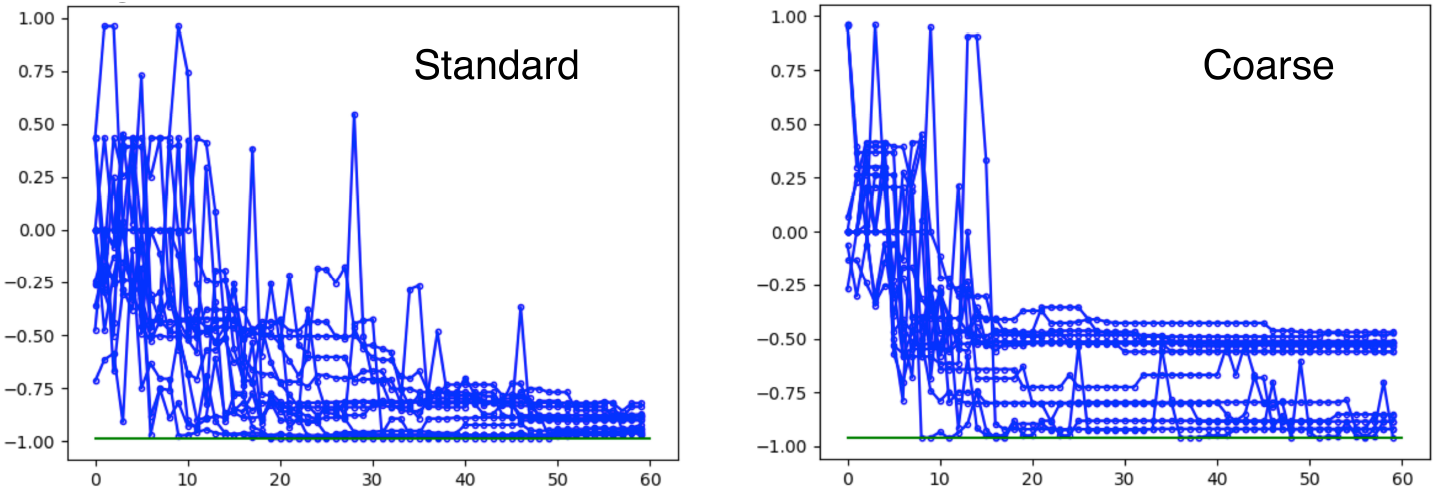

### 3.4 Nonconvexity (local optima)

Our “base” model cost function, as designed for biological plausibility, has the property of *convexity*, i.e. it has a single local minimum which is also the global minimum. In general, optimization is easier for convex functions, and, in particular, functions with multiple local minima are more challenging, because the algorithm can be “trapped” in a local minimum. Therefore, we investigated SafeOpt performance when the cost function was designed to have more than one local minimum.

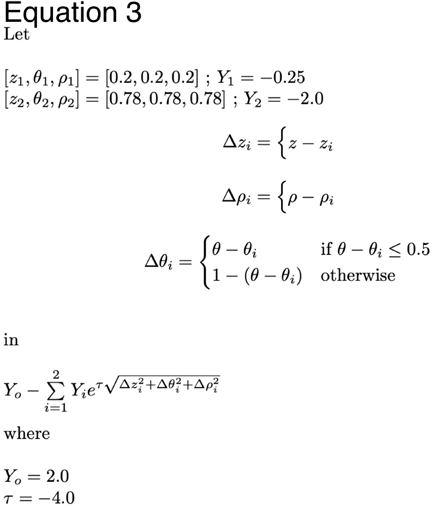

We constructed a cost function with two local minima (Equation 3,Figure 9) It is less smooth than the base function, so we halved the length constant to 0.75. We also found better convergence with a Matern 3/2 kernel, which differs from the Matern 5/2 in allowing smoothness to vary more, from one region to another. Figure 9 shows that the SafeOpt algorithm can “escape” from a local minimum to find the global minimum. <u>*Caption:*</u> True (colormap) and estimated (contour plot) cost function, with test points (white circles) and estimated optimum (red +) at 30,60, and 120 iterations, showing escape from local minimum, and convergance to the global minimum for 16 runs (*lower right)*, though more slowly, and less accurately than in the convex case.

**Figure 9.**
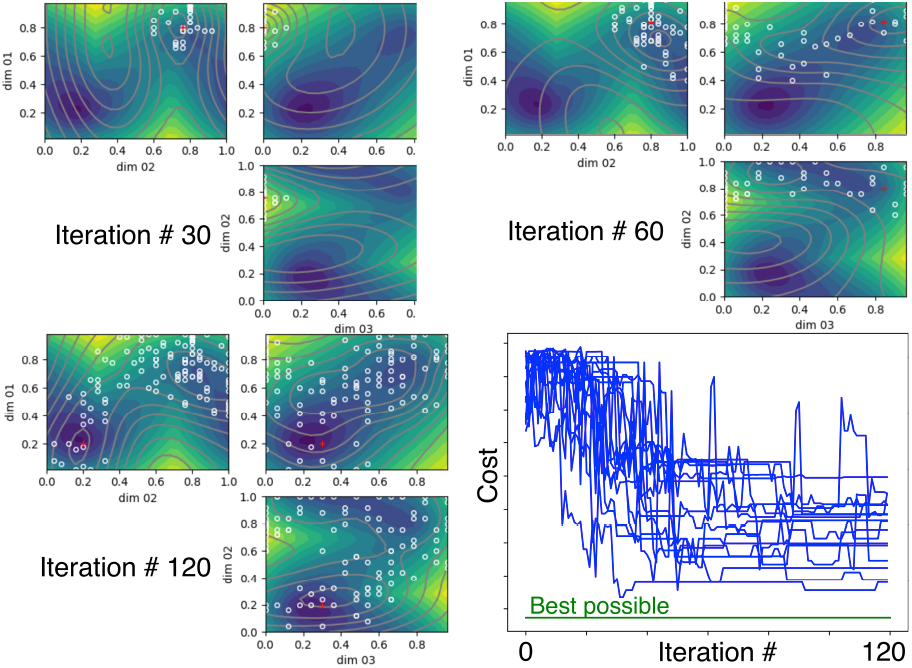

## 4 Discussion

Some symptoms, such as Parkinson’s gait, are often poorly responsive to DBS. This may be because DBS settings are usually optimized to *other* symptoms. To test this, we require an efficient, safe optimization algorithm. Here, in order to develop such a tool, we extend the BayesOpt algorithm whose successful application to DBS settings we previously published^*11*^, using, as a test bed, a simulated cost function constructed for biological plausibility, with measurement noise based on our own experimental data with Parkinson’s gait.

Our extensions are as follows. First, we generalized the algorithm from one to three dimensions. Second, we introduced the SafeOpt algorithm, which, preserves the data efficiency of BayesOpt, but also preserves safety during the learning process. Third, we let the physical properties of the neurostimulator determine the quantization of the stimulation settings, rather than let the algorithm chose values which the neurostimulator could only approximate. Fourth, we allowed a single “known-safe” starting point, rather than starting from randomly placed, possibly unsafe points. (The last two features come naturally with SafeOpt, and we incorporated them into our implementation of BayesOpt, to facilitate a head-to-head comparison, see Appendix). Finally, for the Gaussian process common to both algorithms, we used a kernel specific to the case of a directional DBS electrode, in which one dimension, corresponding to the *direction* stimulation setting, was periodic.

We found that the SafeOpt algorithm converged to the optimum as well, and as fast as the BayesOpt algorithm, while avoiding high-cost points much more effectively. In three dimensions, SafeOpt converged in about 30 iterations, which is a feasible number for physical experiments (i.e. not simulations) in real patients. Consequently, we regard this as our most important finding, since it shows the feasibility of those experiments. Our gait measurement protocol^*19*^ achieved 20 measurements in a single session, and that number could easily have been increased, by eliminating a lengthy set-up procedure which was specific to those experiments. Moreover, the SafeOpt algorithm can be adapted to multiple sessions on different days, with each session resuming where the algorithm left off on the preceding session.

We also explored weaknesses in the algorithm, and what could be done about them. Unsurprisingly, it converged slower when measurement nose was greater, but this could be overcome by running it for more iterations. 60 or even 120 iterations are still feasible, for real patients, particularly if split up over multiple sessions. The algorithm was relatively robust to gross misspecification of multiple hyperparameters, and considerably more robust when hyperparameter fitting was incorporated into the algorithm. The algorithm did not perform as well when the quantization of stimulation settings was coarser, suggesting that it works better with neurostimulators capable of independent current control. Finally, the algorithm was able to cope with a nonconvex cost function (more than one local minimum, of which only one was the global minimum) albeit with an increased number of iterations.

### 4.1 Limitations

The most obvious limitation of our results is that they are based on simulations, rather than physical experiments. Our goal here is to develop an implementation of SafeOpt, and test it in preparation for physical experiments in human patients. To that end, we have used a biologically plausible model cost function, and taken our measurement noise estimate from actual measurements in human patients. We have also explored SafeOpt’s robustness to violation of model assumptions. Importantly, we estimated a required number of iterations in a range that is feasible for human experiments. Without this, we could not consider conducting such experiments. We also verified the superior safety of SafeOpt vs. BayesOpt in our application.

#### 4.1.1 Dimensionality

In this study, we optimized over 3 dimensions (the <u>level</u> *along* the long axis of the electrode; the <u>direction</u> *around* the long axis of the electrode, and the <u>amplitude</u> of stimulation, effectively the *distance from* the electrode to which stimulation extends) which control *where* the electric current is directed, around the electrode. This determines which surrounding neuroanatomical structures are affected: our primary research interest. With three dimensions, the algorithm converged in 30 iterations, but under pessimistic assumptions, took longer, approaching the limits of what would be feasible for experiments in human subjects. Since our goal is to conduct such experiments, we defered simulations in higher dimensions (including frequency and pulse width stimulation parameters) until we have more experimental data.

#### 4.1.2 Convex cost function

We found that both Bayesian optimization methods (SafeOpt and also BayesOpt) performed well if DBS effects can be approximated by a cost function having a single minimum (although performance was also good, with a more complex cost function, if measurement noise was low). This is a natural assumption in the absence of any experimental evidence supporting a more complex cost function. In particular, it is a reasonable assumptison for assessing DBS optimization methods because it is pervasive in the field of clinical DBS research, in which the idea of a “sweet spot” is very widespread^*15*^ i.e. an optimal location within the brain to which electrical current should be directed. Since the three dimensions of stimulation settings in our simulations control the location to which current is directed, the sweet spot concept is appropriate, i.e. a spatial sweet spot corresponds to a sweet spot in the space of stimulation settings. (This differs from the concept of DBS *targets*, i.e. spatially separated brain nuclei such as globus pallidus and subthalamic nucleus, which can be targeted at the time of electrode implantation, but are too far apart to be reached by the same electrode via different stimulation settings.) Although the sweet spot for one symptom may differ from the sweet spot for another, nearly all studies assume that it is unique for a given symptom, apart from a single study (by us^*23*^ in 2014). As noted above, the convexity assumption may be relaxed for symptoms which can be measured with low noise, of which tremor may be an example.

### 4.2 Conclusion

In simulations, the SafeOpt algorithm demonstrated feasibility for physical experiments in real patients

## Data Availability

All data produced in the present study are available upon reasonable request to the authors.

## 5 Appendix: BayesOpt implementation

In our earlier publication^*11*^ we relied on a Matlab library routine for the BayesOpt implementation. For this publication, we reimplemented a version of BayesOpt in Julia. The main reason for this was to facilitate a head to head comparisons, between BayesOpt and SafeOpt with extensive code overlap, including identical GPs. This avoided possible implementation details that could give an “unfair advantage” for one method over the other.

### 5.1 Acquisition function

We used the expected-improvement acquisition function, as in our prior publication. This maximizes the expected value of the decrease in cost, for the proposed test point, relative to the current estimate of the optimum.

### 5.2 Avoiding overexploitation

Expected-improvement tends to overexploit, i.e. it concentrates test points close to the current estimate of the optimum, where model uncertainty is low in preference to exploring high-uncertainty regions, which are more informative, and where a better optimum might lie. Our prior publication ^*11*^ used Matlab’s “expected improvement plus” acquisition function which avoids this by modifying its behavior when it is overexploiting an area. It defines, for a putative test point **x** a measure of overexploitation (σ_x_^2^ - *s*^2^) - (*r*^2^ • *s*^2^) where σ_x_ is the SD of the GP estimate at **x**, *s* is the SD of measurement noise, and *r* is the “exploration ratio” hyperparameter. If this quantity is less than zero, then the point is deemed to be overexploiting, and the algorithm looks for a better, i.e. non-overexploiting test point. In the present paper, as in our previous publication, we use a value of 0.5 for *r*.

When the Matlab BayesOpt generates an overexploiting putative test point, it responds by temporarily increasing the global kernel amplitude parameter and tries again, up to 5 times https://www.math-works.com/matlabcentral/answers/440419. Our implementation of BayesOpt in this paper accomplished this by exploiting the fact that our software framework already classified a finite set of test points as feasible vs. infeasible. We simply classified overexploiting test points as infeasible. This not only maximized code reuse between our BayesOpt and SafeOpt implementation, it also provided a direct guarantee that no non-overexploiting point would be overlooked.

In the case where no non-overexploiting test point could be found, we first tried ignoring the non-overexploiting criterion and simply applying the expected-improvement acquisition function, which is Matlab’s behavior. However, this performed extremely badly: the algorithm overexploited severely, repeatedly choosing test points in the same place, thus gaining very little information about the cost function with each iteration, and failed to find a minimum. Convergence was much better when we altered the behavior to: In the case where no non-overexploiting test point could be found, choose the least-overexploiting test point.

